# Impact of national and regional lockdowns on COVID-19 epidemic waves: Application to the 2020 spring wave in France

**DOI:** 10.1101/2021.04.21.21255876

**Authors:** Jonathan Roux, Clément Massonnaud, Vittoria Colizza, Simon Cauchemez, Pascal Crépey

## Abstract

Several countries have implemented lockdowns to control their COVID-19 epidemic. However, questions like “where” and “when” still require answers. We assessed the impact of national and regional lockdowns considering the French first epidemic wave of COVID-19 as a case study. In a regional lockdown scenario aimed at preventing intensive care units (ICU) saturation, almost all French regions would have had to implement a lockdown within 10 days and 96% of ICU capacities would have been used. For slowly growing epidemics, with a lower reproduction number, the expected delays between regional lockdowns increases. However, the public health costs associated with these delays tend to grow exponentially with time. In a quickly growing pandemic wave, defining the timing of lockdowns at a regional rather than national level delays by a few days the implementation of a nationwide lockdown but leads to substantially higher morbidity, mortality and stress on the healthcare system.

## Introduction

After it was first identified in December 2019, the novel severe acute respiratory syndrome coronavirus 2 (SARS-CoV-2) that causes the 2019 coronavirus disease (COVID-19) quickly spread over the world, challenging health systems. To contain this pandemic, countries implemented control strategies of different intensities for example travel restrictions, closing of schools, shops, stay-at-home orders and lockdowns, at national or regional levels.^1–3^ Combinations of these interventions led to important reductions of the effective reproduction number of the epidemic in a number of countries before the Summer 2020.^1,2,4,5^ France was one of the European countries most affected by the first pandemic wave, along with Italy, Spain and the UK. The first cases of COVID-19 due to importations were confirmed on January 24, 2020.^6^ They were followed by several clusters mostly located in the eastern regions of France (Grand-Est and Bourgogne-Franche-Comté) and Ile-de-France (the most populated French region including the city of Paris). The seeding of the epidemic in these regions mean that they were ahead of other regions in terms of the number of COVID-19 patients admitted into intensive care units (ICU) (mean of 53 and 31 daily ICU admissions from March 11 to March 17 in Ile-de-France and Grand-Est, respectively, compared to 1 to 12 in other French regions).^5,7^ To avoid saturation of the healthcare system, the French government ordered a national lockdown starting on March 17, 2020.

The efficacy of lockdown strategies to control COVID-19 epidemics has been internationally demonstrated at the national ^1,2,5,8,9^ and county levels.^10^ However, some have argued that the nationwide application in France of such measures was unnecessary and that lockdowns restricted to the two or three most impacted French regions would have been sufficient to contain the pandemic wave. The present study aimed at retrospectively comparing the impact of the nationwide lockdown on March 17 to regional lockdowns on the number of COVID-19 hospitalizations, occupied ICU beds, life-years gained, and deaths avoided during the first pandemic wave in France.

## Results

### Pre-lockdown and lockdown epidemiological dynamics

Table 1 presents estimates of the reproduction number before R_prelockdown_ and during the lockdown period R_lockdown_ by region (see Supplementary Figures S1 to S4 for the fitting of hospital admissions, ICU admissions, occupied ICU beds and deaths). The median regional reproduction number before the national lockdown was 2.60 with a minimum of 1.94 (95% confidence interval [1.76-2.15]) in Grand-Est and a maximum of 4.17 [3.39-5.10] in Centre-Val de Loire (Table 1). After the implementation of the lockdown, the median reproduction number was estimated at 0.71. The lockdown was implemented on March 17, 2020 at a time when there were 6.50 daily hospital admissions per 100 000 inhabitants in the Grand-Est region (reference region). Using the pre-lockdown reproduction number, we found that the date when this threshold would have been reached without lockdown ranged from March 18 for Ile-de-France to April 7 for Bretagne (Table 1, GES threshold). Using the same process, we found that all regions but Bretagne would have implemented a lockdown by March 27, 2020 in order to avoid ICU saturation (Table 1, ICU capacity threshold).

**Table 1.** Estimated values of R_prelockdown_ and R_lockdown_ and dates when GES threshold and ICU capacity threshold would have been reached.

| Region | R before lockdown<br>[95%CI] | R during lockdown<br>[95%CI] | GES threshold date | ICU capacity<br>threshold date |
| --- | --- | --- | --- | --- |
| GES | 1.94 [1.76-2.15] | 0.78 [0.68-0.88] | March 17 | March 19 |
| IDF | 2.41 [2.24-2.60] | 0.73 [0.66-0.80] | March 18 | March 14 |
| BFC | 2.60 [2.28-2.96] | 0.71 [0.60-0.82] | March 25 | March 17 |
| ARA | 2.97 [2.65-3.32] | 0.69 [0.60-0.78] | March 26 | March 20 |
| CVL | 4.17 [3.40-5.10] | 0.75 [0.60-0.94] | March 26 | March 18 |
| HDF | 1.96 [1.79-2.14] | 0.73 [0.65-0.81] | March 27 | March 21 |
| PAC | 2.32 [2.07-2.60] | 0.74 [0.64-0.84] | March 28 | March 26 |
| PDL | 3.55 [2.94-4.26] | 0.68 [0.55-0.84] | March 29 | March 24 |
| NOR | 2.96 [2.55-3.44] | 0.66 [0.55-0.78] | March 31 | March 24 |
| OCC | 2.79 [2.44-3.19] | 0.64 [0.54-0.74] | March 31 | March 25 |
| NAQ | 3.29 [2.75-3.92] | 0.67 [0.55-0.82] | April 2 | March 27 |
| BRE | 2.26 [1.85-2.74] | 0.69 [0.54-0.85] | April 7 | April 2 |

95%CI: 95% Confidence Interval. GES threshold date: The date on which the estimated incidence of hospital admissions reached 6.50 per 100 000 inhabitants in the region. ICU capacity threshold date: The last date on which a lockdown would allow the region to stay below its ICU capacity limit.
GES: Grand-Est, IDF: Ile-de-France, BFC: Bourgogne-Franche-Comté, ARA: Auvergne-Rhône-Alpes, CVL: Centre-Val de Loire, HDF: Hauts de France, PAC: Provence-Alpes-Côte d'Azur, PDL: Pays de la Loire, NOR: Normandie, OCC: Occitanie, NAQ: Nouvelle-Aquitaine, BRE: Bretagne.

### Impact of a national lockdown

At the national level, setting a national lockdown one day later than the observed lockdown, i.e on March 18, would have led to an increase of 1,449 ICU admissions, the need for 829 additional ICU beds, and 1,765 deaths (i.e. 2 ICU admissions and 3 deaths per 100,000 inhabitants); corresponding to a loss of almost 16,200 QALY (Figure 1 – regional results are presented in Supplementary Tables S1 and S2 and Supplementary Figure S5). It would have led to an increase in the maximum number of occupied ICU beds at the peak between 8% and 20% at the regional level (Figure 2); in hospital admissions between 7% and 18%; in ICU admissions between 7% and 18%; and in deaths between 7% and 18% (Supplementary Figures S6 to S8). A national lockdown one week after the official start, i.e. on March 24, would have led to 12,868 additional ICU admissions and 15,822 additional deaths (i.e. 20 ICU admissions and 25 deaths per 100 000 inhabitants); corresponding to 144,000 QALY lost. Moreover, 15,668 ICU beds would have been needed to accommodate all ICU admissions and would have led to an overrun of ICU capacity with 145% occupancy of ICU beds (Figure 1 and 2).

**Figure 1.**
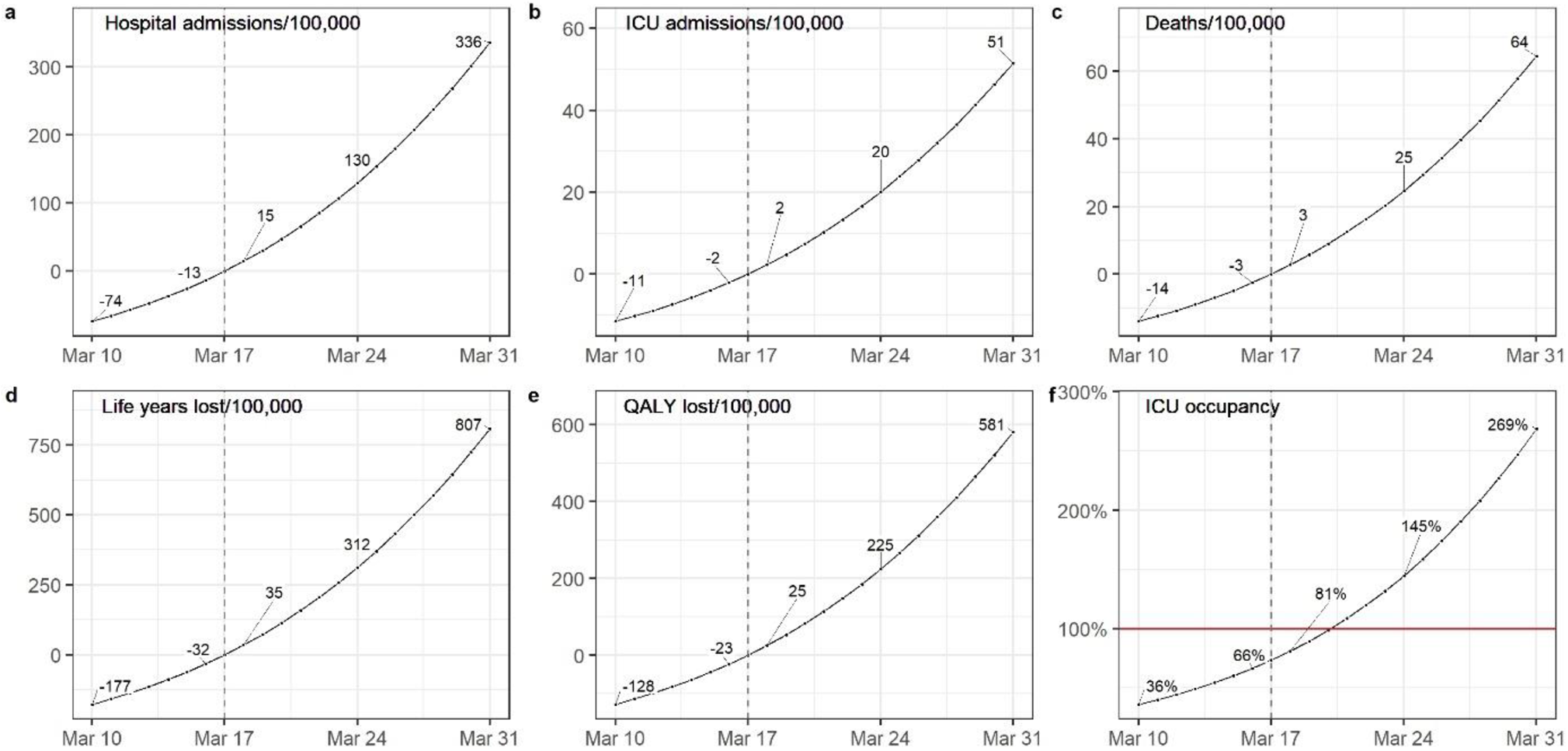
Impact of the change of the lockdown date on the national number of (a) hospital admissions, (b) ICU admissions, (c) deaths, (d) life years, (e) quality-adjusted life years lost per 100 000 inhabitants compared to the observed lockdown date and on (f) the ICU occupancy. ICU: Intensive care unit, QALY: Quality-adjusted life years.

**Figure 2.**
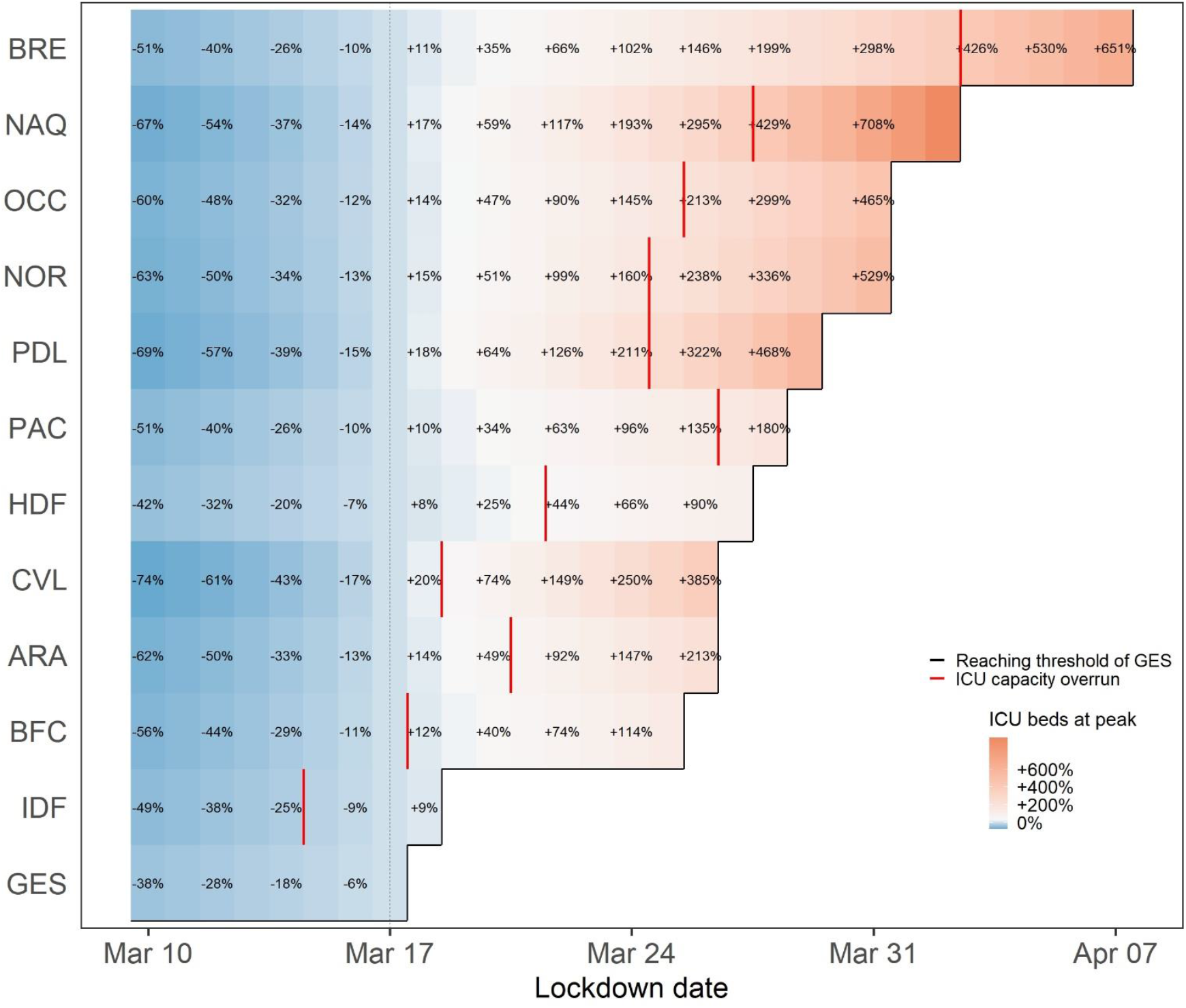
Relative change in the number of occupied ICU beds at the peak for each French metropolitan region (in %) according to lockdown date compared to the observed national lockdown. ICU: Intensive care unit, GES: Grand-Est, IDF: Ile-de-France, BFC: Bourgogne-Franche-Comté, ARA: Auvergne-Rhône-Alpes, CVL: Centre-Val de Loire, HDF: Hauts de France, PAC: Provence-Alpes-Côte d’Azur, PDL: Pays de la Loire, NOR: Normandie, OCC: Occitanie, NAQ: Nouvelle-Aquitaine, BRE: Bretagne.

Starting the lockdown one day earlier, i.e. on March 16 instead of March 17, would have led to 1,337 fewer ICU admissions and 1,625 fewer deaths, corresponding to around 14,900 QALY saved. In addition, 7,189 ICU beds would have been needed at peak, corresponding to 756 fewer ICU beds than what was observed. Locally, it would have led to a reduction in the range 6%-16% for the number of deaths and 7-16% for the number of ICU admissions (Supplementary Figures S7 and S8). If the global lockdown had started 7 days earlier on March 10, it would have led to 7,369 fewer ICU admissions, 8,925 fewer deaths and around 82,000 QALY saved. Moreover, 3,913 fewer ICU beds would have been needed at peak, corresponding to approximately half the number of ICU beds used at the peak of the wave.

### Impact of regional lockdowns

The first scenario, where the start of the lockdown is delayed in each region according to the GES threshold (the date on which the estimated incidence of hospital admissions reached 6.50 per 100,000 inhabitants in the region), would have led to 17,409 additional ICU admissions, 20,719 additional deaths and the need for 16,671 ICU beds at the national level. Approximately 262,000 life-years and 188,800 QALY would have been lost with this delay. It would have resulted in a peak of daily hospital admissions ranging from 405 in Bretagne to 1,663 in Ile-de-France (Supplementary Figure S9). This scenario would have led to the saturation of ICU capacities in some regions (Figure 2). In the scenario where each region implements a lockdown no later than the moment allowing them to remain within its ICU capacity (ICU capacity threshold), Ile-de-France would have had to be locked down before the start of the observed national lockdown on March 17, and one region (Bretagne) could have been locked down three weeks later on April 2, 2020. However, all regions but Bretagne would have had to implement a lockdown by March 27, 2020. By this date, a maximum of 95.6% of national ICU capacities would have been used, instead of less than 70% with the observed national lockdown 10 days earlier, and almost 6,000 more lives would have been lost (Figure 3).

**Figure 3.**
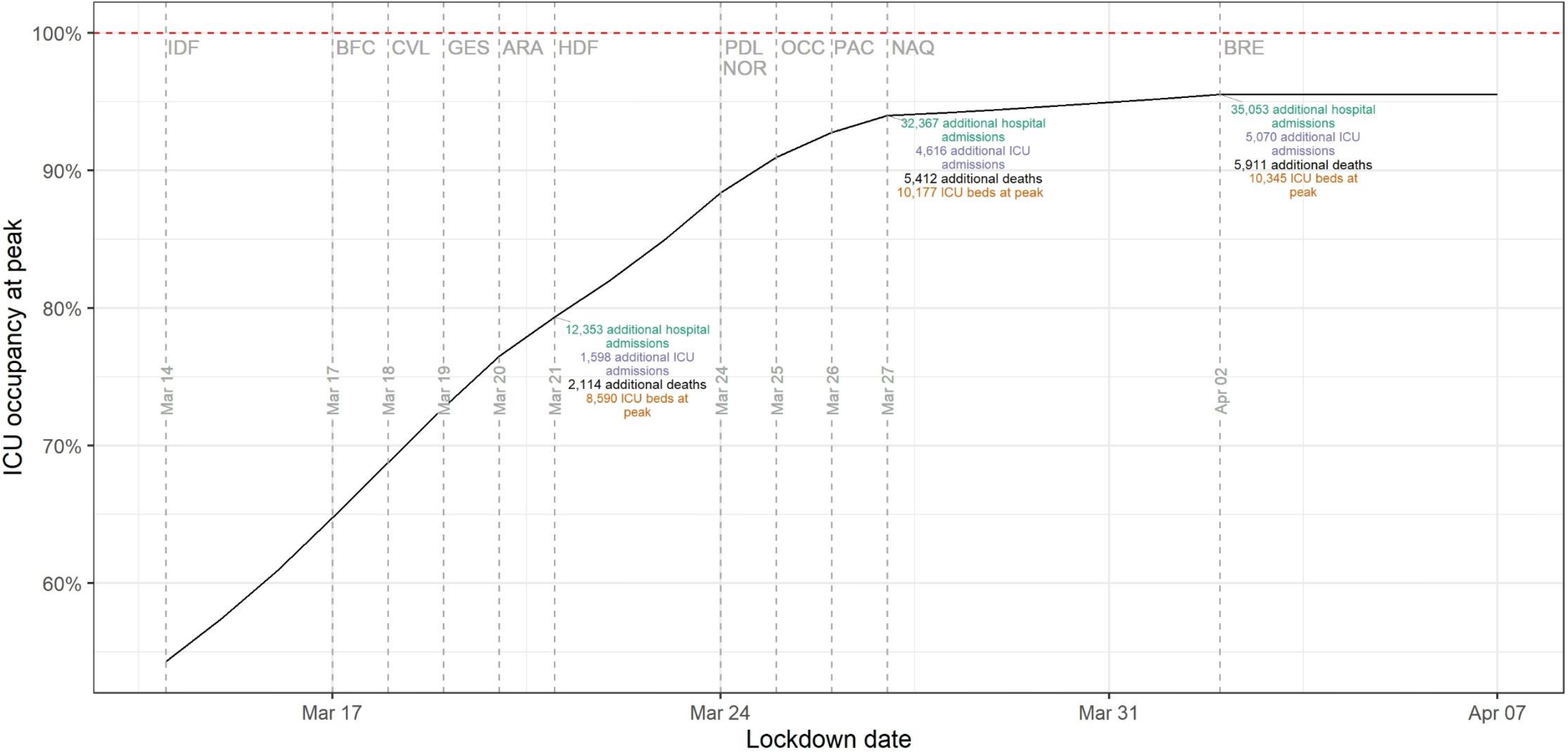
ICU beds occupancy in case of regional lockdowns to avoid regional ICU saturation. Vertical dashed lines indicate the moment when regions have to be locked down to avoid a saturation of their ICU capacity. Three regions (GES, IDF, BFC) would be locked from the start of the study period. One region (BRE) could have been locked on April 2, 2020, all others would have been locked before March 27, 2020. ICU: Intensive care unit, GES: Grand-Est, IDF: Ile-de-France, BFC: Bourgogne-Franche-Comté, ARA: Auvergne-Rhône-Alpes, CVL: Centre-Val de Loire, HDF: Hauts de France, PAC: Provence-Alpes-Côte d’Azur, PDL: Pays de la Loire, NOR: Normandie, OCC: Occitanie, NAQ: Nouvelle-Aquitaine, BRE: Bretagne.

### Scenario with a slower epidemic growth

The sensitivity analysis showed that the lockdown impact depends not only on timing but also on the pre-lockdown reproduction number R_prelockdown_ observed in the region (Supplementary Figure S10). For example, delaying the national lockdown two weeks later on March 31 would have led to 1,334 additional deaths with R_prelockdown_=1.1 but 5,913 additional deaths with R_prelockdown_=1.5. In addition, for any given reproduction number, the cost of delaying the lockdown increases exponentially with time, for all outcomes. As an example, a synchronized lockdown on March 10 with a reproduction number equal to 1.5 would have led to 2,105 fewer deaths, whereas on March 24 it would have led to 2,653 additional deaths.

## Discussion

This study aimed at comparing the impact of lockdown strategies where the timing of the lockdown is defined at the national vs at the regional scale, in the context of a quickly growing pandemic wave. This was done thanks to a compartmental model that reproduced the epidemic dynamics of the Spring 2020 pandemic wave in France and simulated several counterfactual scenarios to that of the nationwide lockdown implemented on March 17.

Using our model, we estimated that, at the time of the first national lockdown, all regions were not at the same epidemic stage. Indeed, the threshold of 6.50 hospital admissions per 100,000 inhabitants estimated in the most affected region at the time lockdown took place (Grand-Est region), would have been reached in other regions within 1 day (Ile-de-France) - 21 days (Bretagne) after March 17, in the absence of lockdown. As expected, delaying the lockdown would have led to additional hospital and ICU admissions, and deaths, independently of the region. Waiting to reach this threshold in each region would have resulted in an increase of 17,409 ICU admissions and 20,719 deaths at the national level, comparable to the total number of ICU admissions and deaths registered in the country until July 1.^7^ This delay would have resulted in a 8-fold increase of deaths in some regions (Nouvelle-Aquitaine) compared to the observed outcome. Moreover, these numbers do not take into account the additional deaths due to healthcare system congestion. Therefore, it is likely that the lockdown would have been implemented before the saturation of all regions. On the other hand, starting the national lockdown one week earlier would have resulted into up to 8,925 fewer deaths, corresponding to around 82,000 QALY lost at the national level. The reduction was higher in regions where the reproduction number was high, highlighting the importance of this parameter in the decision to start a lockdown.

We also analysed the impact of regional lockdowns. Using a threshold based on the number of hospital admissions to trigger a regional lockdown failed to prevent regions from exceeding their ICU capacity. This is easily explained by different ICU capacities per inhabitant by region, and because the impact of lockdown timing depends not only on the incidence but also on the epidemic dynamics, described by the reproduction number R. Depending on R, the same level of incidence in two regions on a given day can lead to very different dynamics in the following days, as shown in the scenario with a slower epidemic growth. The higher the reproduction number, the higher the cost of delaying the lockdown. Our simulations showed that regional lockdowns, where each lockdown is started on the last date that allows the region to stay within its ICU capacity, would have led to 6,000 additional deaths compared to the observed lockdown. In this scenario, all regions but one would have been locked down within 10 days. Although each region would have stayed within its ICU capacities, 96% of national ICU capacities would have been used at peak. These assessments are performed retrospectively. In real-time, it would be difficult to precisely estimate the date when the lockdown should be implemented to avoid the saturation of local ICUs. As a consequence, there is a real risk that the regional approach would have led to the saturation of ICUs in some locations. Considering the latest information on SARS-CoV-2 and its characteristics, our estimated values of reproduction number were similar to the ones obtained by other researchers before and after the lockdown.^4,11,12^ Regarding the computation of outcomes of interest (hospital requirements and deaths), we considered the whole care pathway of patients inside hospital settings (Figure S13). In addition, all these estimations were based on observed parameters (lengths of stay, ICU admission risk, and death risk) in real settings and modulated for each region with specific estimated coefficients to account for regional disparities. However, some differences may remain, mainly due to patients’ transfers or to change in hospitalization strategies during the crisis. Concerning the quantification of the impact of a delayed lockdown, we chose Grand-Est as the reference region, as it was the first impacted region of the first wave of the epidemic in France, and thus the number of hospitalizations in this region at lockdown start was a good threshold for comparisons.^5^

The mortality estimates presented in this study do not consider deaths occurring outside hospitals (at home – estimated to be around 5% of all COVID-related deaths^13^ – or in nursing homes –10,497 deaths recorded as of July 2, 2020)^14^ as we limited our analysis to hospital settings. Moreover, our model also does not account for the excess mortality that would have resulted from hospital saturation. Given the important excess hospital and ICU admissions that would have occurred with a delayed lockdown, it is likely that the saturation of the health care system would have resulted in an important excess mortality. This was avoided in France thanks to the transfer of 658 patients from highly affected to less affected regions, which remained less infected thanks to the national lockdown (personal communication). Therefore, our analysis may underestimate the number of deaths, life years and QALY lost in scenarios where delays in the implementation of the lockdown increase substantially the stress on the healthcare system. In addition, QALY lost were only estimated based on the deaths of infected individuals, whereas it would be interesting to take into account the lost QALY due to severe COVID-19 leading to hospitalization and ICU admissions, since there may still be after-effects for these individuals.^15^ However, to our knowledge, no study has been done in France to provide utility measures in these cases. Regarding the estimation of ICU occupancy, the maximum ICU capacity in each region corresponded to the maximum number of beds that could be mobilised during the pandemic, which was higher than the normal number of ICU beds at the national level. Without this mobilisation of new ICU beds, ICU saturation would have been reached much earlier. Finally, our study relied on the hypothesis that implementing regionals lockdowns is doable and acceptable by the population. Some European countries such as Italy,^3,16^ Germany^3^ or United Kingdom^17^ have implemented such measures. In France, spatially targeted measures were introduced only after the first wave and for less restrictive measures than a full lockdown.^18^ On one hand it allows to limit the implementation of strong control measures in areas where the infection risk is high, and potentially acknowledged by the population. On the other hand, it raises questions regarding the consequences related to population movements that may occur following or preceding the lockdown of a region.^10^ In addition, some studies have shown that localized mitigation measures, less strict than national lockdown (curfew in specific metropolitan area, closing of shops or schools), have led to a reduction in the French population mobility^19^ and a slowdown of the epidemic in non-restricted areas.^18^

This study highlights that the impact of a delayed regional lockdown greatly depends on the reproduction number in the region, with costs in terms of morbi-mortality growing exponentially with the delay. It also shows that in a quickly growing pandemic wave such as the one observed in Spring 2020 in France, defining the timing of lockdowns at a regional rather than national level delays by only a few days the implementation of a nationwide lockdown but leads to substantially higher morbi-mortality and stress on the healthcare system.

## Methods

### Transmission model

We used a deterministic, age-structured, compartmental epidemic model based on demographic and age profile of the population of the 13 administrative regions of metropolitan France (Supplementary Figure S11). We considered that individuals were susceptible (S), and then potentially exposed to the virus but not infectious (E). As observed in clinical practice, we set children and adolescents to be less susceptible to infection than other age groups.^20^ Exposed individuals stay in their compartment for an average of 2.72 days^21^ before moving to either asymptomatic (A) or pre-symptomatic (I_ps_) compartment according to observed risk of being asymptomatic.^20^ We considered that asymptomatic individuals were 45% less infectious than pre-symptomatic individuals were^22^ and we assumed they stayed in the compartment for an average of 10.91 days before moving to the removed compartment (R) of patients that are cured or dead from COVID-19. Pre-symptomatic individuals will become infected symptomatic (I_s_) after an average duration of 2.38 days. This choice of parameters gave a mean incubation period of 5.1 days,^1^ within around 2 days of pre-symptomatic transmissions. Then they become after an average of 5 days (assumption) either hospitalized (I_h_) or remain within the community (I_nh_) according to age-dependent hospitalization risks^23,24^ adjusted to apply only to clinical cases.^20^ We assumed that hospitalized individuals were no more infectious because of their hospitalization whereas non-hospitalized keeps spreading the disease with the same intensity as symptomatic individuals. We assumed the average duration in I_nh_ compartment was 3.53 days to match the total duration in the asymptomatic compartment. Finally, both hospitalized and non-hospitalized individuals moved to the removed compartment.

To match the epidemic dynamics and reproduce the Erlang distributions of durations in each compartment of the transmission model, we subdivided each compartments having a role in the infection process into sub-compartments. This subdivision had no impact on the mean duration spent in each compartment. A sensitivity analysis testing several combinations revealed that 10 sub-compartments in each compartment permitted to approach the observed dynamics of the epidemic (Supplementary Figure S12).

### Model parametrization

The population was divided into 17 age groups: 16 age-band of 5 years from 0 to 80 years, and a last group for people aged 80 years and older. The population structure of each region was inferred from hospital catchment areas from 2016 and 2017 census data provided by the French National Institute of Statistics and Economic Studies (Insee).^25,26^ To simulate age and location-dependent mixing, we used inter-individual contacts matrices for the French population estimated by Prem et al.^27^We retrieved epidemiological regional data related to the COVID-19 epidemic in metropolitan France gathered by the French National Public Health Agency (SpF-’Santé publique France’)^7^ daily number of hospital admissions (general and intensive care unit (ICU) wards), daily number of ICU admissions, daily number of occupied ICU beds, and daily number of deaths in hospitals (deaths in nursing homes and at home were not considered). All these epidemiological data were corrected for reporting delays following the same procedure as Salje et al.^4^ We also obtained data on the maximum ICU beds capacity per French region from the ‘Direction de la Recherche, des Études, de l’Évaluation et des Statistiques’ (Drees) regarding the situation during the March 2020 lockdown (personal communication) (Supplementary Table S3).

### Estimation of hospital-related outcomes

Based on the estimated number of new infected hospitalized cases per day provided by our epidemiological model, we inferred outcomes related to hospital requirements, namely ICU admissions, ICU occupied beds, and deaths.

We divided the hospital settings in two parts: general ward and ICU ward (Supplementary Figure S13). The epidemiological transmission model estimated the daily number of new hospitalized cases due to COVID-19 infection, regardless of the ward (i.e. ICU and general wards). Once admitted to hospital, infected cases could either remain in the general ward until the end of their stay or go into ICU, if they became severe cases. We assumed that cases admitted in ICU entered ICU ward the same day as they were admitted in hospital (pre-ICU length of stay equal to 0 day). Once in ICU, cases could either die or stay in ICU until their discharge to general ward. Cases in general ward could either die or stay in general ward until their discharge to home.

We used age-dependent ICU admission risks for hospitalized patients estimated by Salje et al.^4^ We also used age-specific lengths of stay in ICU to estimate the number of occupied ICU beds.^28^ We estimated the number of deaths using hospital and ICU death risks estimated by the Drees on all the hospitalized cases of the first wave of epidemic in France (March-June 2020).^28^ Deaths were delayed in time using the time from hospital or ICU admission to death.^28^ Lengths of stay in general ward before discharge, lengths of stay in general ward before death and post-ICU lengths of stay in general ward were not used as we did not estimated the total number of hospital beds needed. This had no impact on the results provided by the model.

We also estimated the number of life years and quality-adjusted life years (QALY) lost for each death using life tables provided by INSEE for 2012-2016^29^ and utility measures of each age-group in France.^30^

### Statistical framework

We estimated region-specific model parameters by maximum likelihood in a two-step process using the *bbmle* R package.^31^ First, on the period stretching from March 14 to May 10, 2020, corresponding to the evolution of the epidemic until the end of the national lockdown, we estimated the value of the transmission parameter β, governing the value of R_0_, the initial state in each compartment per age group on March 1, 2020 and the effects of the national lockdown. The latter was estimated through a transmission reduction parameter, hereafter called *c*_*β*_, and multiplied the transmission parameter (and thus contact matrices) to reproduce the several mitigation measures implemented and their consequences on the regional propagation. We jointly estimated these parameters by fitting the daily de-seasonalized time series of hospital admissions (hereafter denoted Hosp) using the likelihood *L*_*β*_ defined in Equation 1.

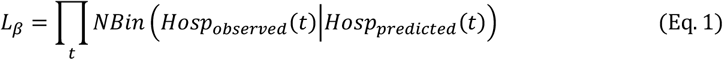

where NBin(.|X) is a negative binomial distribution of mean X and overdispersion *X*^*δ*^, *δ* being a parameter specific to each region to be estimated. Confidence intervals of these two parameters were estimated using likelihood profiling methods.^31^

In a second step, we jointly estimated three regional coefficients adjusting age-specific risks of ICU admissions, risk of deaths and lengths of stay in ICU (including ICU stays leading to death). They were estimated by simultaneously fitting the time series of ICU admissions (hereafter denoted ICU), deaths (both smoothed using 7-day centered moving average) and occupied ICU beds (hereafter denoted BedICU) from March 14 to May 10, 2020 using the likelihood *L*_*ICU*−*deaths*_ defined in Equation 2.

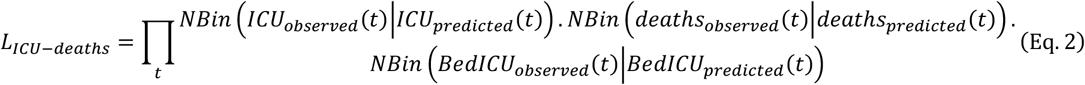

where NBin(.|X) is a negative binomial distribution of mean X and overdispersion *X*^*δ*^, *δ* being a parameter estimated for each region.

### Analysis of national and regional lockdowns

For each region, we estimated the reproduction number before the lockdown (R_prelockdown_), and after its implementation (R_lockdown_). Then, in order to simulate a regional lockdown, we used the R_prelockdown_ up to the starting date of the measure; and the R_lockdown_ estimated in the region after that date. We assumed that the transmission rate remained constant until July 1. We also assumed that the epidemic dynamic was driven by local behaviour within regions and that interactions between regions had no impact on local reproduction numbers.

We simulated national lockdowns starting on all dates from March 10 to March 31, thus exploring other dates of start beyond the actual date of March 17. We then simulated asynchronous regional lockdowns, in which each region could be locked down independently of other regions, on different dates. We considered the following epidemic thresholds as starting dates of regional lockdowns: either (1) the date on which the estimated incidence of hospital admissions in the region reached the level of the most affected region during the first wave (i.e. Grand-Est, here called GES threshold), or (2) the last date on which a lockdown would allow the region to stay below its ICU capacity limit (here called ICU capacity threshold).

### Outcomes

For each simulation, we computed at the national and regional levels the cumulative numbers of hospital and ICU admissions, maximum occupied ICU beds, deaths, life years and quality-adjusted life years (QALY) lost between March 1 and July 1 and computed the relative change compared to the outcomes obtained when the real lockdown date was used. The transmission model was implemented in C++. Data collection, data management, simulations, results analysis and reporting were performed using R.^32^

### Scenario with a slower epidemic growth

To better understand how the reproduction number affect the estimated impact of the lockdown, we analysed scenarios where R_prelockdown_ is in the range 1.1 to 1.5. This corresponds to the reproduction numbers observed in France at the start of 2021. In this analysis, we kept the previously estimated R_lockdown_ for each region during the lockdown.

## Data Availability

The data and code underlying this article will be shared on reasonable request to the corresponding author.

## Data availability

The data that support the findings of this study are available from the corresponding author upon request.

## Acknowledgements

This work was supported by the French national research agency (ANR), through the SPHINx (Spread of Pathogens on Healthcare Institutions Networks) project [grant number SPHINX-17-CE36-0008-01].

## Author contributions

J.R. and P.C. conceived the project, designed and conducted the analyses, interpreted the results, and wrote the first draft of the manuscript; C.M., V.C. and S.M. discussed the results and contributed to revisions of the manuscript. All authors contributed to the final version of the paper and approved the final manuscript.

## Additional Information

## Competing interests

The authors declare no competing interests.

**Correspondence** and requests for materials should be addressed to P.C. or J.R.

## Supplementary information

**Supplementary Table S1.**
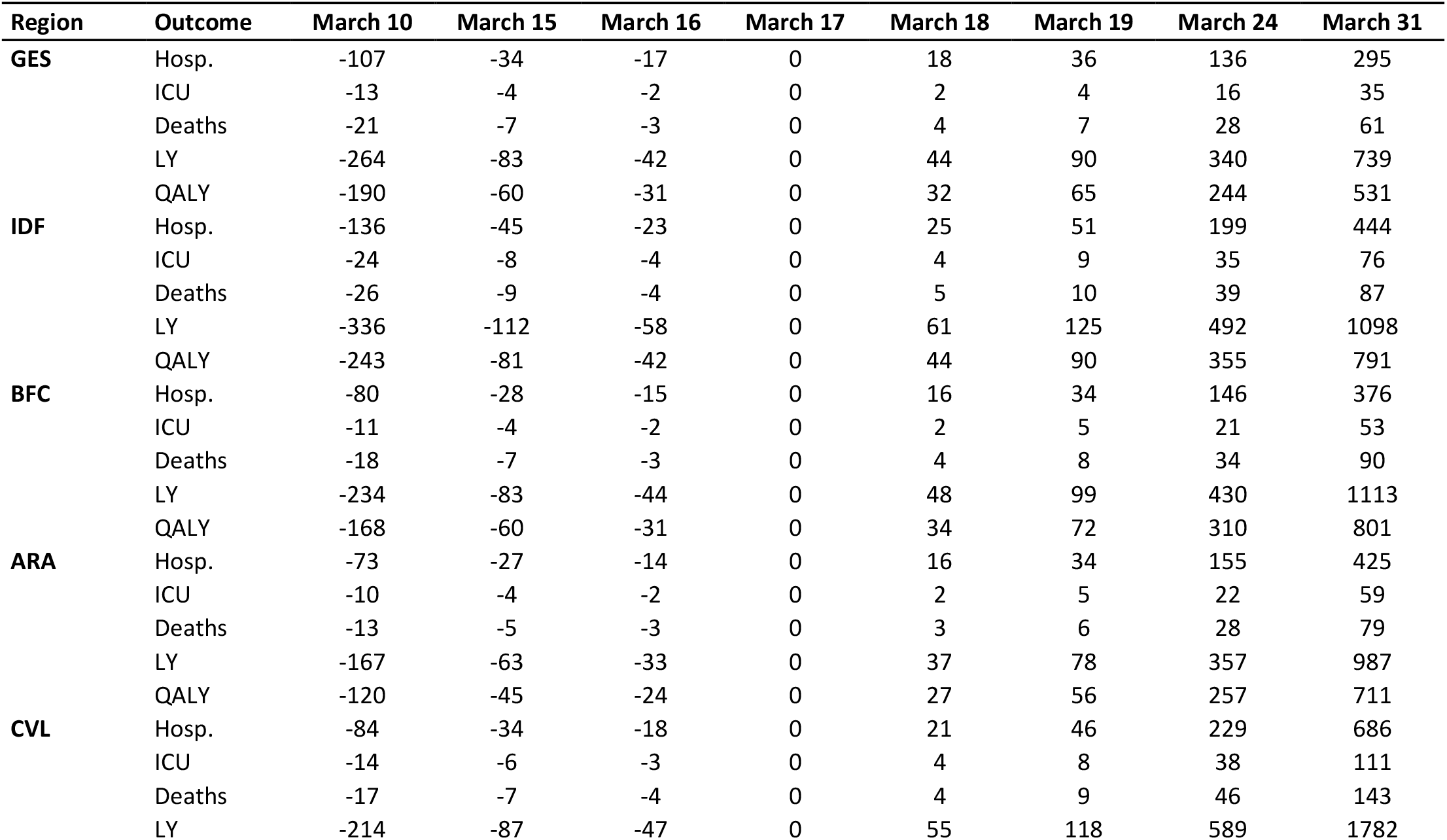
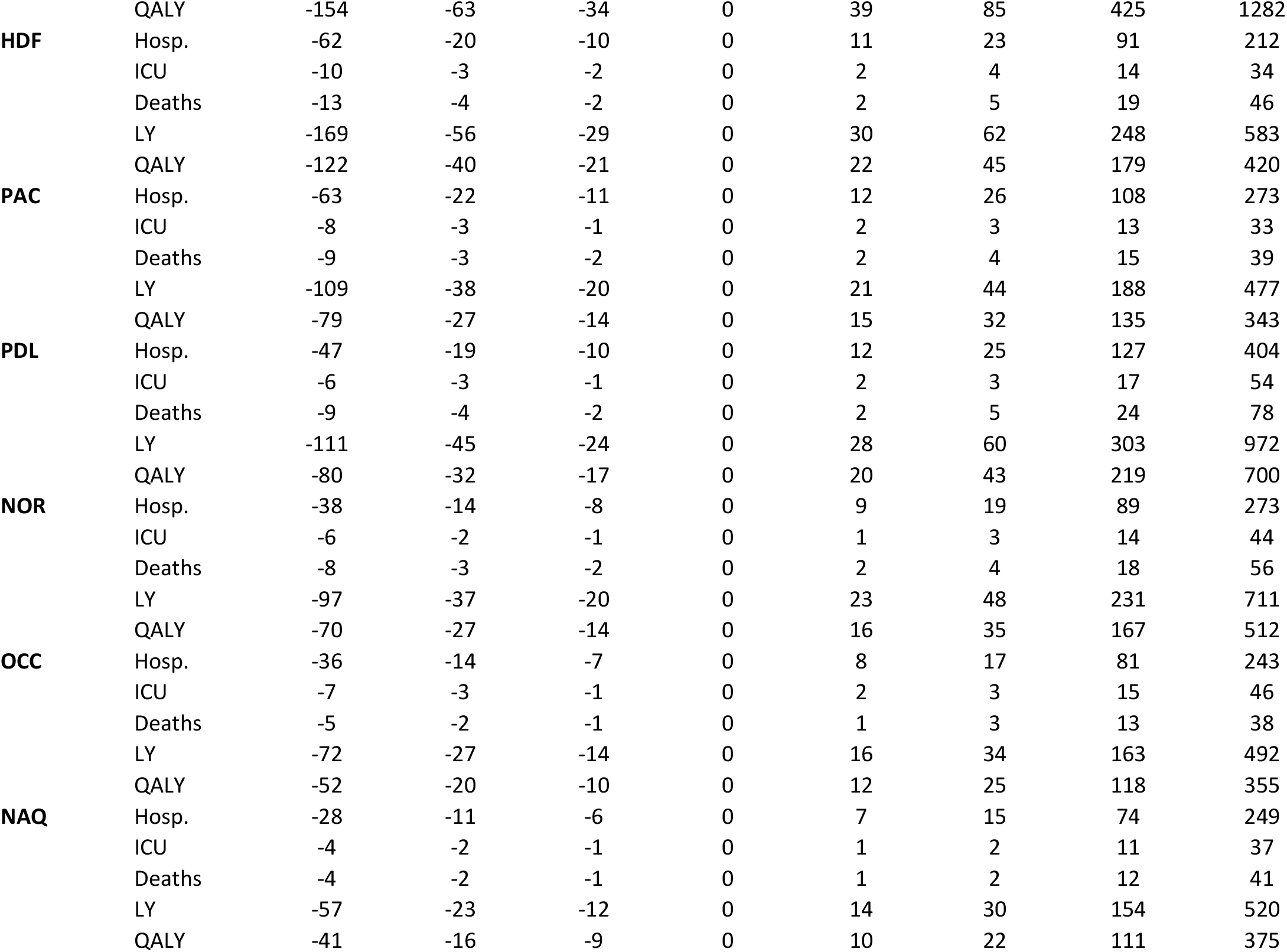
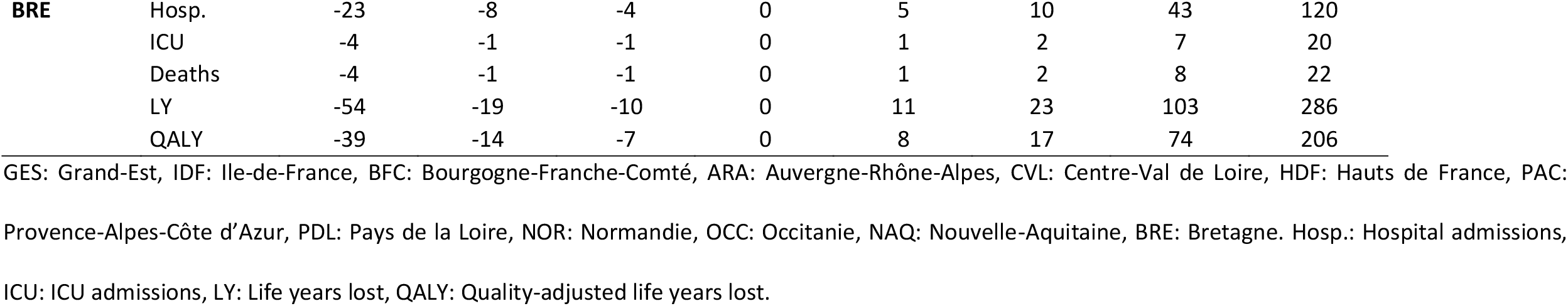
Impact of the change of the lockdown date on the number of hospital and ICU admissions, deaths, life years lost and quality-adjusted life years lost per 100,000 inhabitants compared to the observed lockdown date per French Region.

**Supplementary Table S2.** Impact of the change of the lockdown date on the regional occupation of ICU beds and the maximum ICU beds needed at the peak per French region

| Region | Outcome | March 10 | March 15 | March 16 | March 17 | March 18 | March 19 | March 24 | March 31 |
| --- | --- | --- | --- | --- | --- | --- | --- | --- | --- |
| **GES** | Occupancy (%) | 54 | 76 | 81 | 87 | 92 | 98 | 132 | 187 |
|  | Max ICU beds | 657 | 926 | 990 | 1056 | 1126 | 1198 | 1609 | 2276 |
| **IDF** | Occupancy (%) | 67 | 109 | 120 | 131 | 144 | 157 | 237 | 376 |
|  | Max ICU beds | 1920 | 3141 | 3452 | 3786 | 4144 | 4530 | 6823 | 10834 |
| **BFC** | Occupancy (%) | 43 | 77 | 87 | 97 | 109 | 122 | 209 | 406 |
|  | Max ICU beds | 146 | 262 | 294 | 330 | 370 | 413 | 707 | 1377 |
| **ARA** | Occupancy (%) | 23 | 47 | 53 | 61 | 70 | 80 | 150 | 330 |
|  | Max ICU beds | 268 | 535 | 613 | 701 | 801 | 915 | 1729 | 3792 |
| **CVL** | Occupancy (%) | 20 | 52 | 62 | 75 | 91 | 109 | 264 | 748 |
|  | Max ICU beds | 53 | 139 | 168 | 203 | 245 | 295 | 712 | 2020 |
| **HDF** | Occupancy (%) | 41 | 61 | 66 | 71 | 77 | 82 | 118 | 186 |
|  | Max ICU beds | 342 | 505 | 545 | 588 | 633 | 682 | 975 | 1536 |
| **PAC** | Occupancy (%) | 20 | 33 | 37 | 41 | 45 | 50 | 80 | 146 |
|  | Max ICU beds | 196 | 326 | 361 | 399 | 440 | 486 | 782 | 1434 |
| **PDL** | Occupancy (%) | 10 | 22 | 26 | 31 | 37 | 43 | 97 | 269 |
|  | Max ICU beds | 44 | 102 | 120 | 142 | 168 | 198 | 441 | 1228 |
| **NOR** | Occupancy (%) | 13 | 26 | 30 | 34 | 39 | 45 | 89 | 216 |
|  | Max ICU beds | 63 | 127 | 147 | 169 | 194 | 222 | 438 | 1061 |
| **OCC** | Occupancy (%) | 13 | 25 | 28 | 32 | 37 | 42 | 79 | 183 |
|  | Max ICU beds | 124 | 241 | 274 | 313 | 356 | 405 | 765 | 1766 |
| **NAQ** | Occupancy (%) | 7 | 15 | 18 | 21 | 25 | 29 | 62 | 171 |
|  | Max ICU beds | 60 | 131 | 153 | 179 | 209 | 244 | 524 | 1444 |
| **BRE** | Occupancy (%) | 10 | 17 | 18 | 20 | 22 | 25 | 41 | 81 |
|  | Max ICU beds | 39 | 66 | 73 | 80 | 89 | 98 | 162 | 320 |

GES: Grand-Est, IDF: Ile-de-France, BFC: Bourgogne-Franche-Comté, ARA: Auvergne-Rhône-Alpes, CVL: Centre-Val de Loire, HDF: Hauts de France, PAC:
Provence-Alpes-Côte d'Azur, PDL: Pays de la Loire, NOR: Normandie, OCC: Occitanie, NAQ: Nouvelle-Aquitaine, BRE: Bretagne. ICU : Intensive care unit.
Occupancy: Occupancy of ICU beds. Max ICU beds: Maximum number of ICU beds needed.

**Supplementary Table S3.** Maximum number of ICU beds available per region in March 2020

| Region | Maximum number of ICU beds |
| --- | --- |
| GES | 1219 |
| IDF | 2885 |
| BFC | 339 |
| ARA | 1150 |
| CVL | 270 |
| HDF | 827 |
| PAC | 980 |
| PDL | 456 |
| NOR | 492 |
| OCC | 967 |
| NAQ | 844 |
| BRE | 397 |
| France | 10 826 |

ICU : Intensive care unit. GES: Grand-Est, IDF: Ile-de-France, BFC: Bourgogne-Franche-Comté, ARA: Auvergne-Rhône-Alpes, CVL: Centre-Val de Loire, HDF: Hauts de France, PAC: Provence-Alpes-Côte d'Azur, PDL: Pays de la Loire, NOR: Normandie, OCC: Occitanie, NAQ: Nouvelle-Aquitaine, BRE: Bretagne.

**Supplementary Figure S1.**
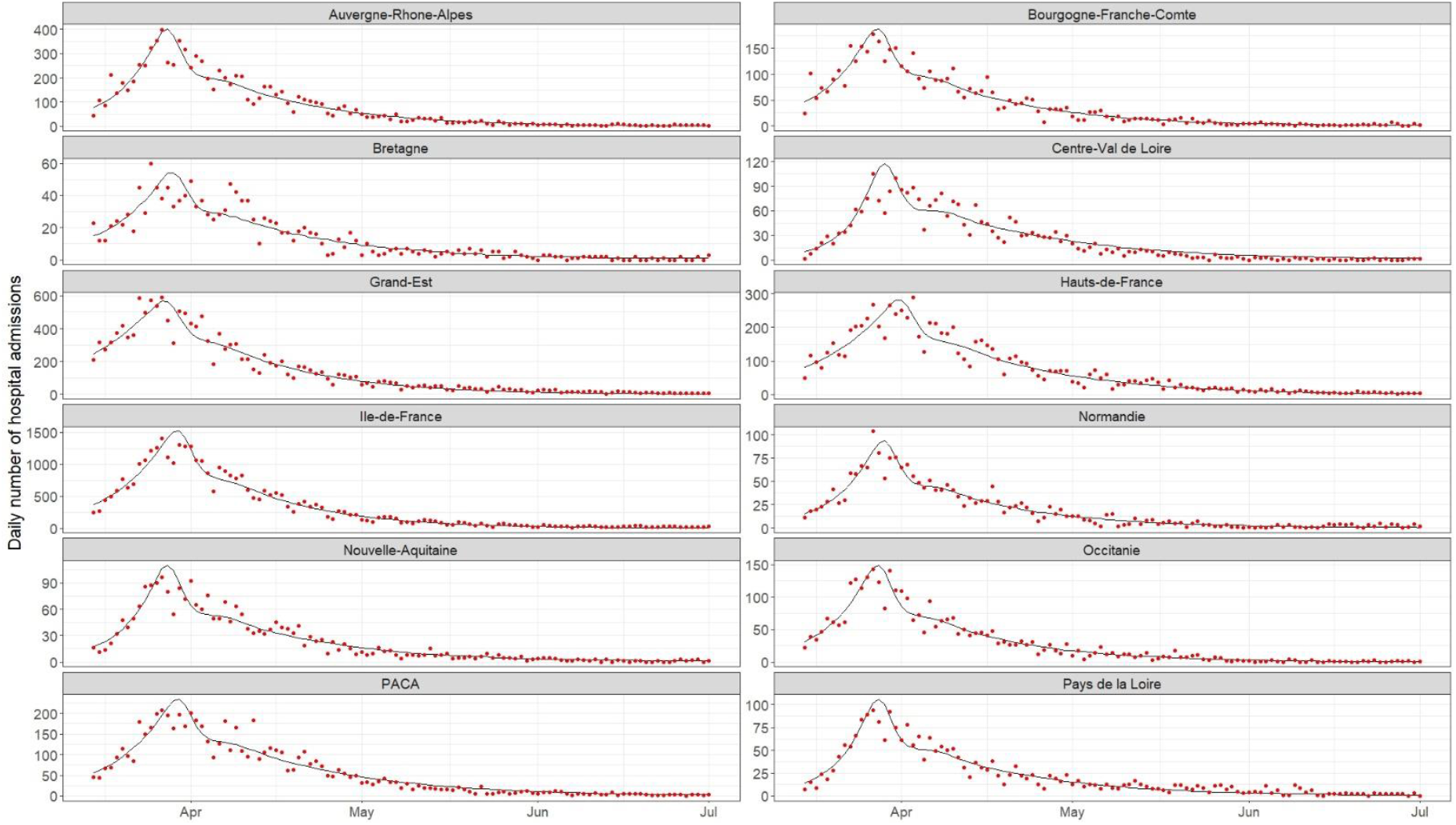
Prediction of the number of new hospital admissions per French region between March 15 and July 1, 2020. The black line stands for the predicted values and the red dots for the observed values.

**Supplementary Figure S2.**
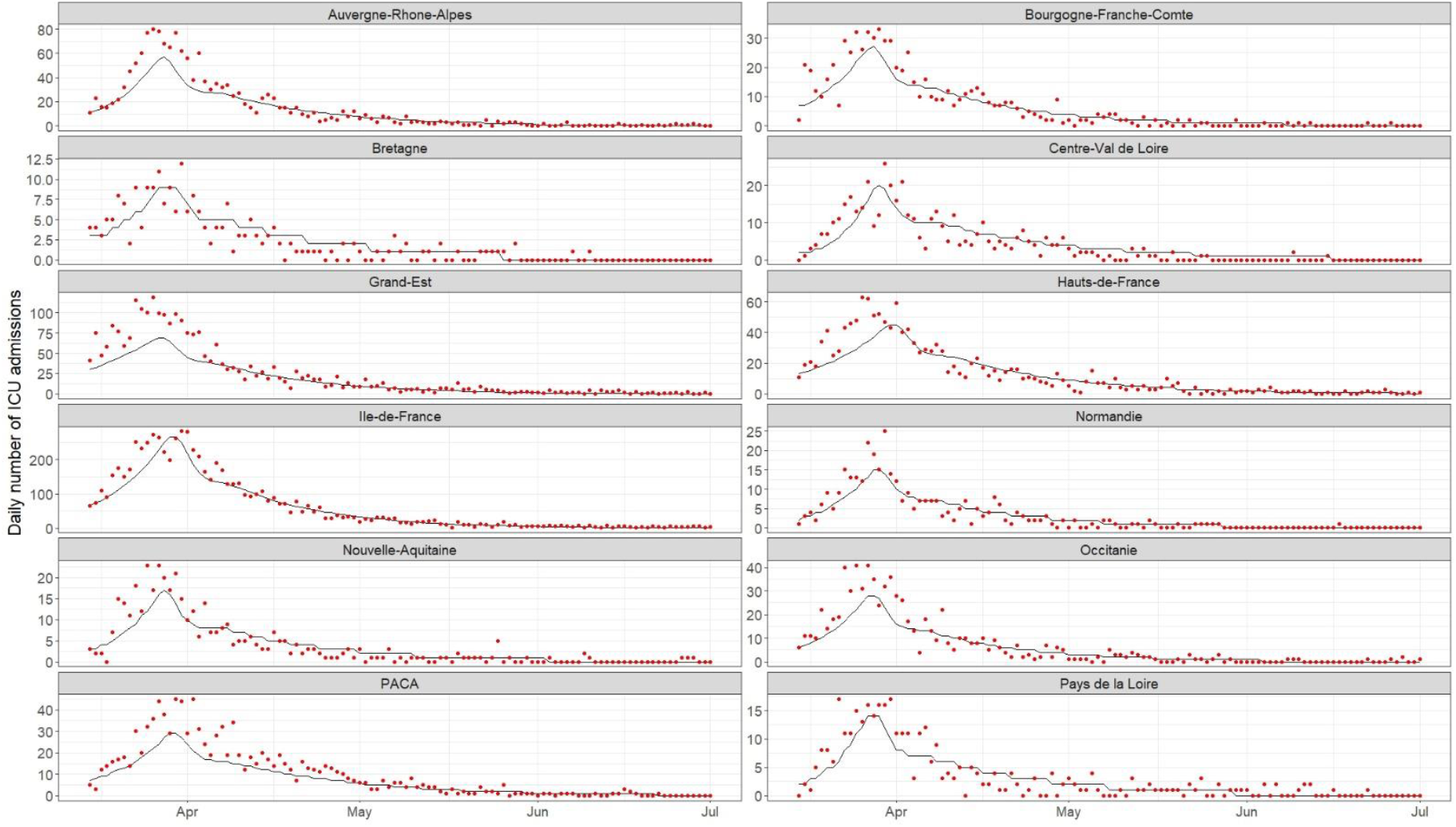
Prediction of the number of new ICU admissions per French region between March 15 and July 1, 2020. The black line stands for the predicted values and the red dots for the observed values. ICU: Intensive care unit.

**Supplementary Figure S3.**
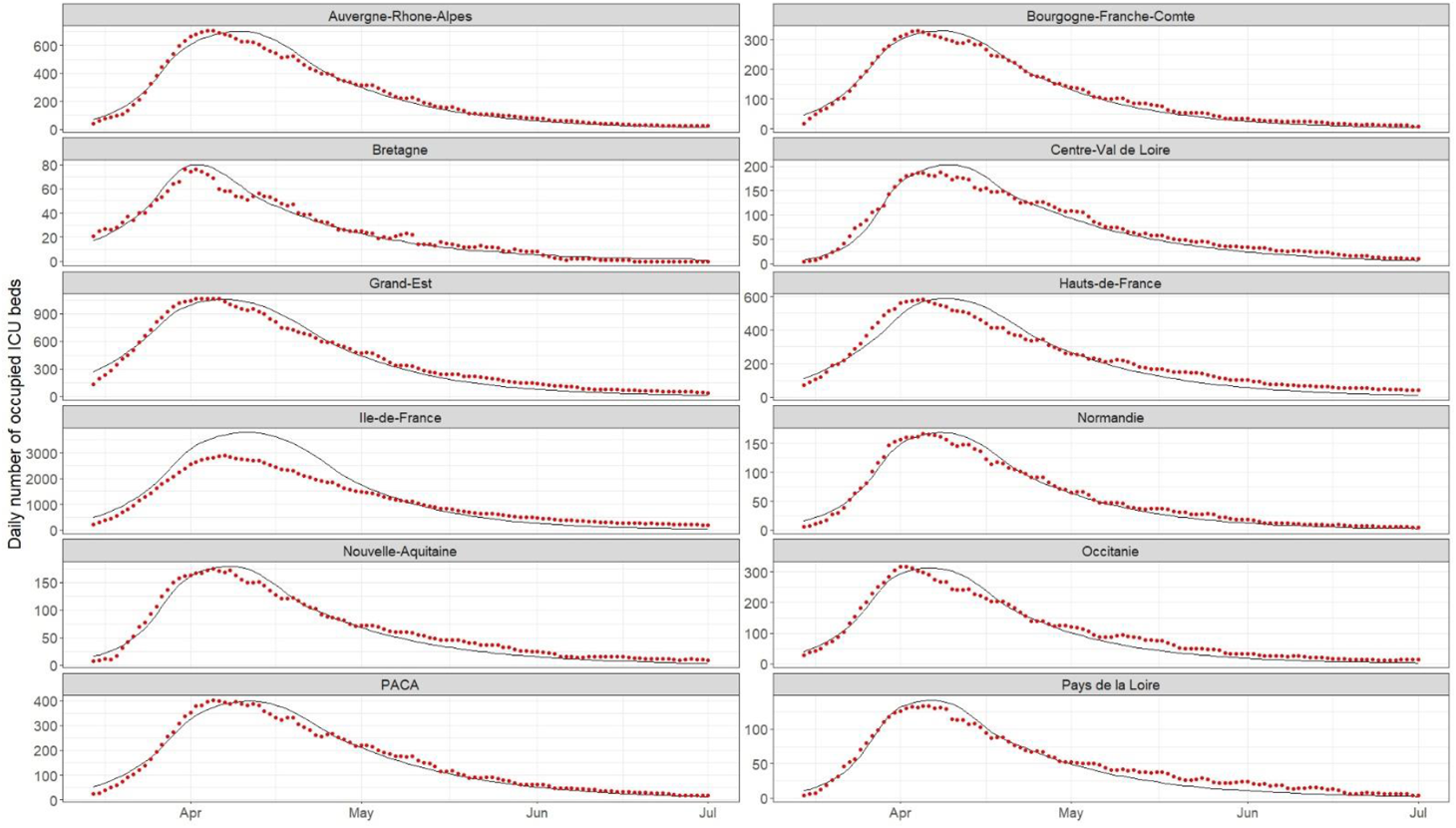
Prediction of the number of occupied ICU beds per French region between March 15 and July 1, 2020. The black line stands for the predicted values and the red dots for the observed values. ICU: Intensive care unit.

**Supplementary Figure S4.**
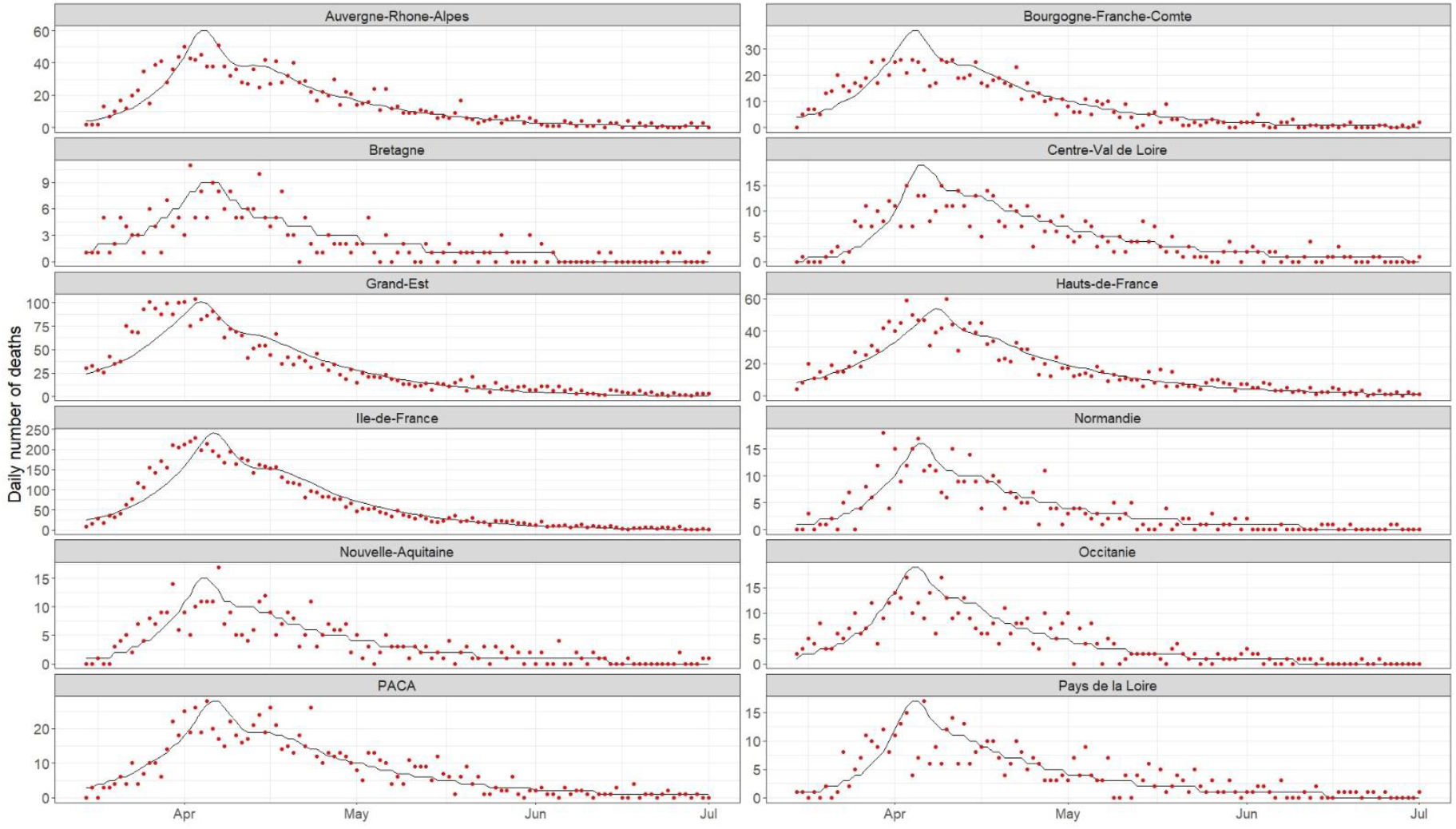
Prediction of the number of new deaths per French region between March 15 and July 1, 2020. The black line stands for the predicted values and the red dots for the observed values.

**Supplementary Figure S5.**
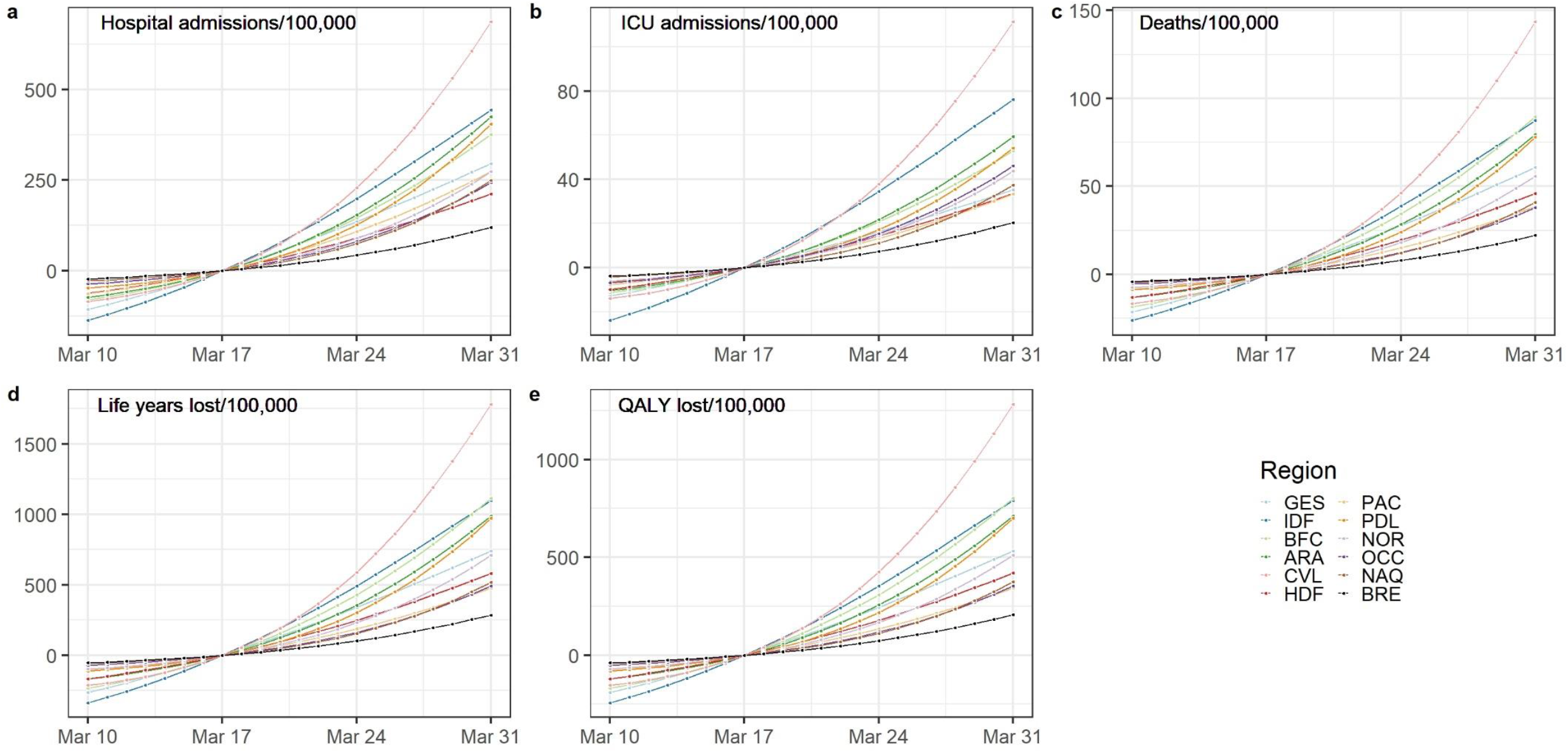
Impact of the change of the lockdown date on the regional number of (a) hospital and (b) ICU admissions, (c) deaths, (d) life years and (e) quality-adjusted life years lost per 100,000 inhabitants compared to the observed lockdown date. ICU: Intensive care unit. QALY : Quality-adjusted life years. GES: Grand-Est, IDF: Ile-de-France, BFC: Bourgogne-Franche-Comté, ARA: Auvergne-Rhône-Alpes, CVL: Centre-Val de Loire, HDF: Hauts de France, PAC: Provence-Alpes-Côte d’Azur, PDL: Pays de la Loire, NOR: Normandie, OCC: Occitanie, NAQ: Nouvelle-Aquitaine, BRE: Bretagne.

**Supplementary Figure S6.**
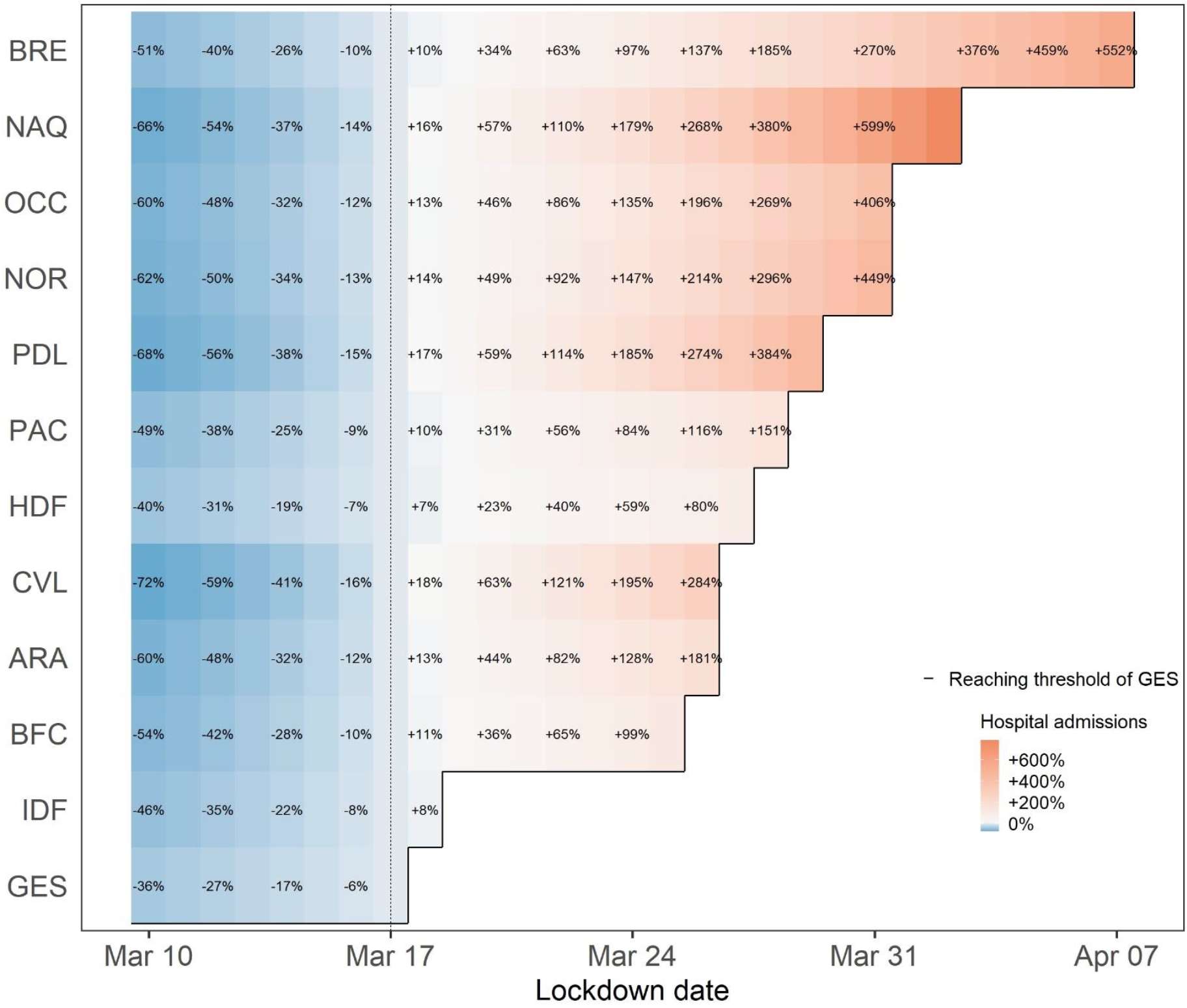
Relative change in the number of hospital admissions (in %) according to lockdown date compared to the real lockdown for each French metropolitan region. GES: Grand-Est, IDF: Ile-de-France, BFC: Bourgogne-Franche-Comté, ARA: Auvergne-Rhône-Alpes, CVL: Centre-Val de Loire, HDF: Hauts de France, PAC: Provence-Alpes-Côte d’Azur, PDL: Pays de la Loire, NOR: Normandie, OCC: Occitanie, NAQ: Nouvelle-Aquitaine, BRE: Bretagne.

**Supplementary Figure S7.**
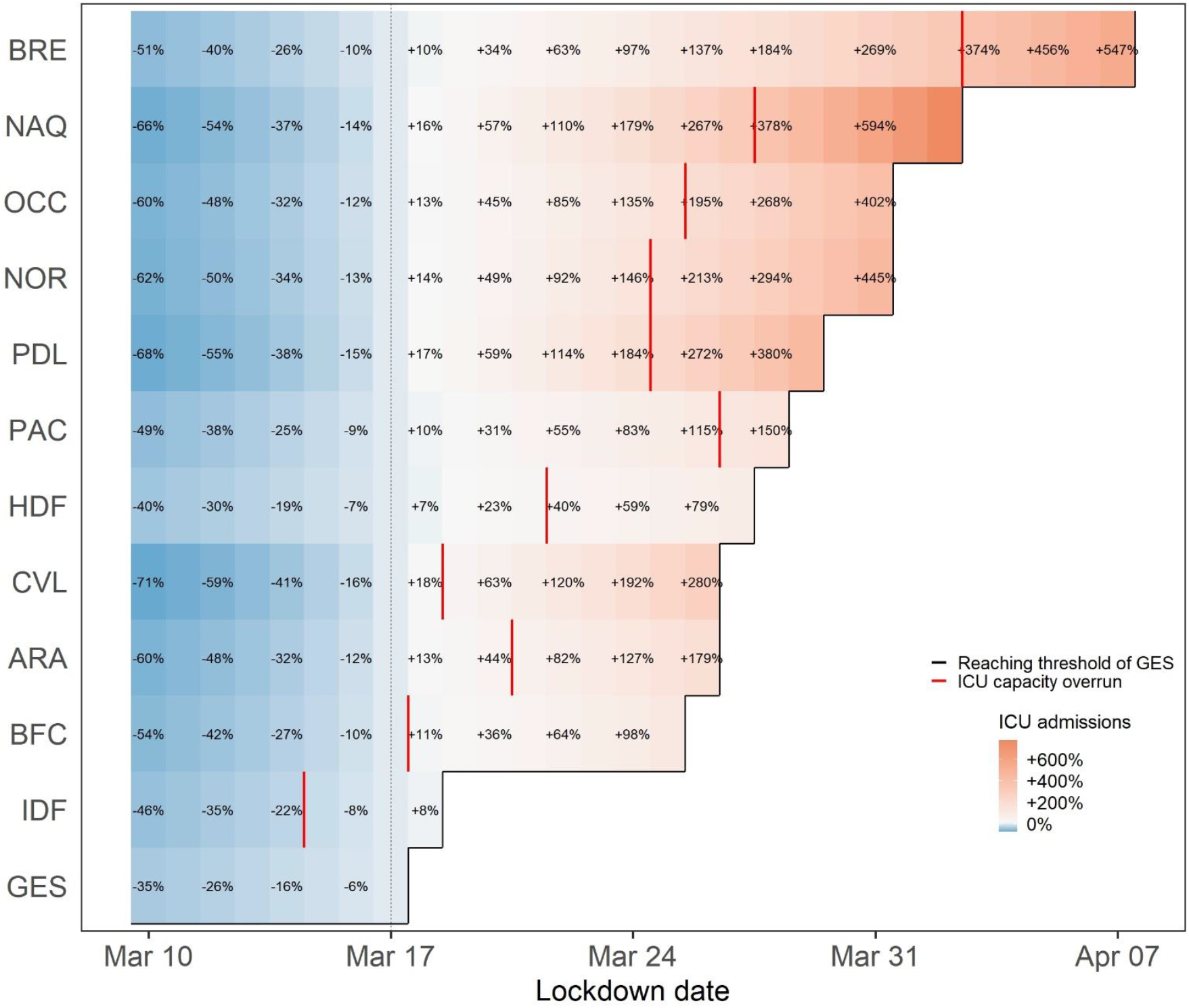
Relative change in the number of ICU admissions (in %) according to lockdown date compared to the real lockdown for each French metropolitan region. ICU: Intensive care unit. GES: Grand-Est, IDF: Ile-de-France, BFC: Bourgogne-Franche-Comté, ARA: Auvergne-Rhône-Alpes, CVL: Centre-Val de Loire, HDF: Hauts de France, PAC: Provence-Alpes-Côte d’Azur, PDL: Pays de la Loire, NOR: Normandie, OCC: Occitanie, NAQ: Nouvelle-Aquitaine, BRE: Bretagne.

**Supplementary Figure S8.**
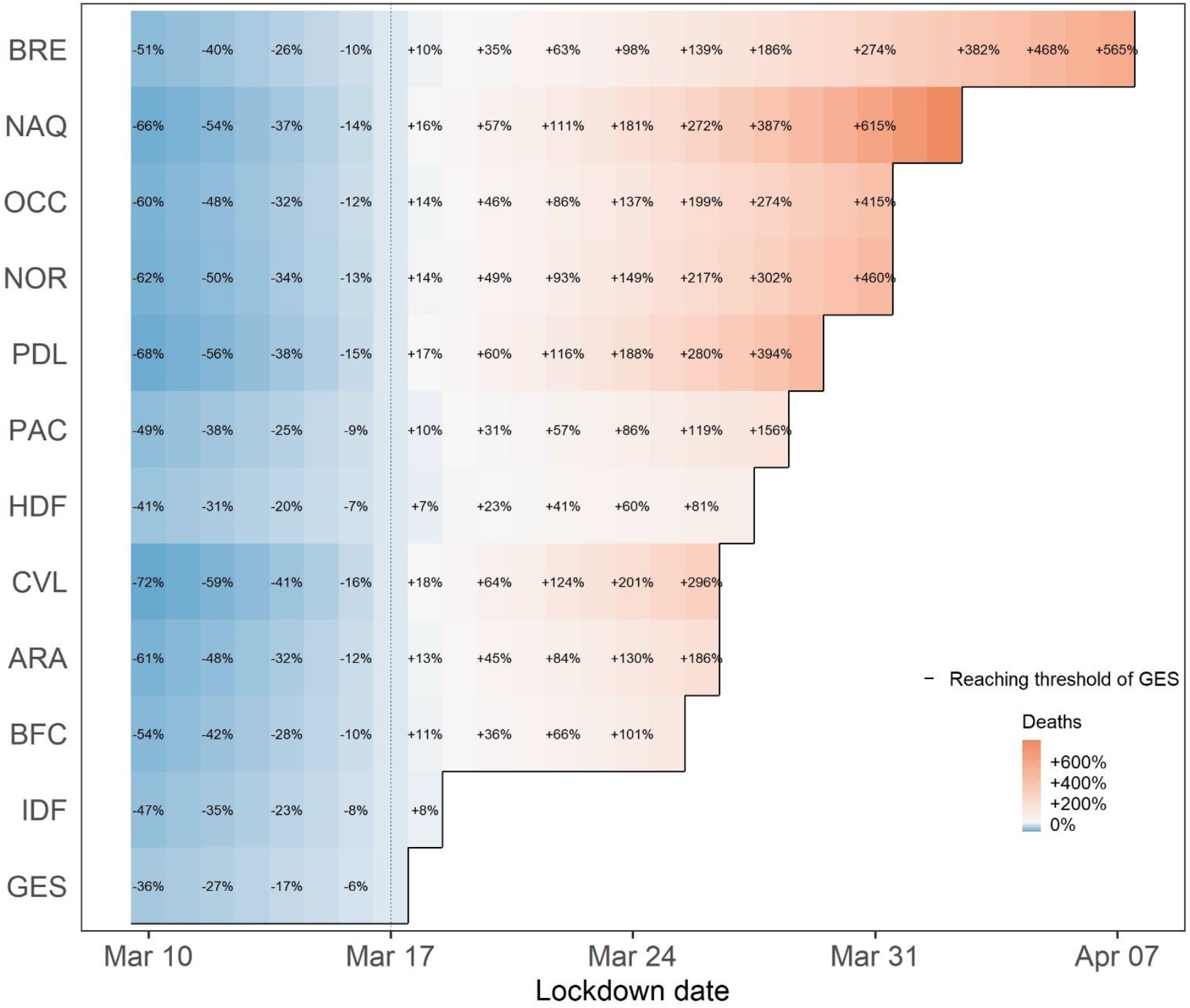
Relative change in the number of deaths (in %) according to lockdown date compared to the real lockdown for each French metropolitan region. GES: Grand-Est, IDF: Ile-de-France, BFC: Bourgogne-Franche-Comté, ARA: Auvergne-Rhône-Alpes, CVL: Centre-Val de Loire, HDF: Hauts de France, PAC: Provence-Alpes-Côte d’Azur, PDL: Pays de la Loire, NOR: Normandie, OCC: Occitanie, NAQ: Nouvelle-Aquitaine, BRE: Bretagne.

**Supplementary Figure S9.**
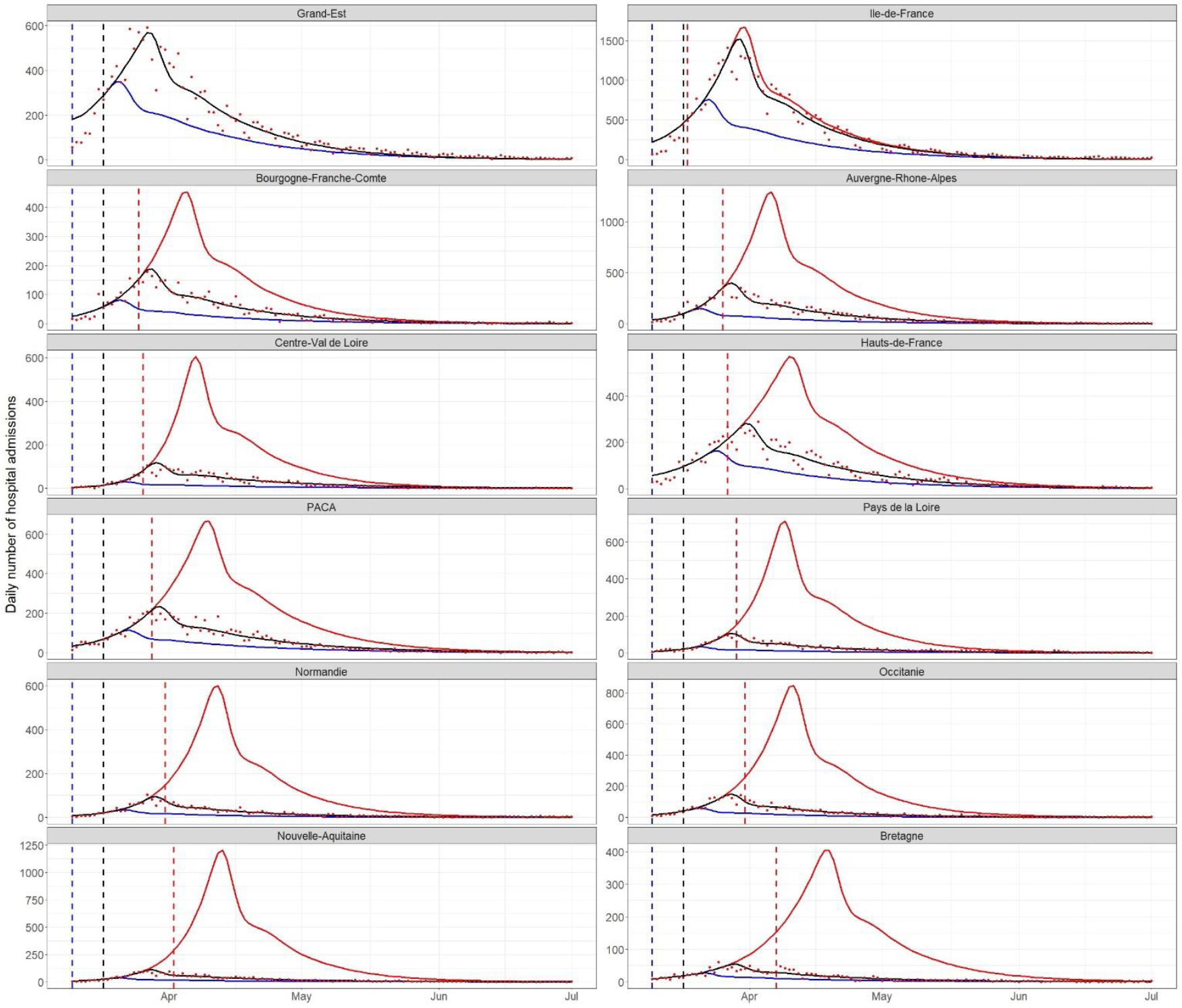
Prediction of the number of new hospital admissions per French region between March 10 and July 1, 2020 with a lockdown start on March 10 (blue), March 17 (black) and GES threshold date (the date on which the estimated incidence of hospital admissions per 100,000 inhabitants in the region reached 6.50) (red). The dashed lines correspond to the starting dates of each lockdown and the red points to the observed values.

**Supplementary Figure S10.**
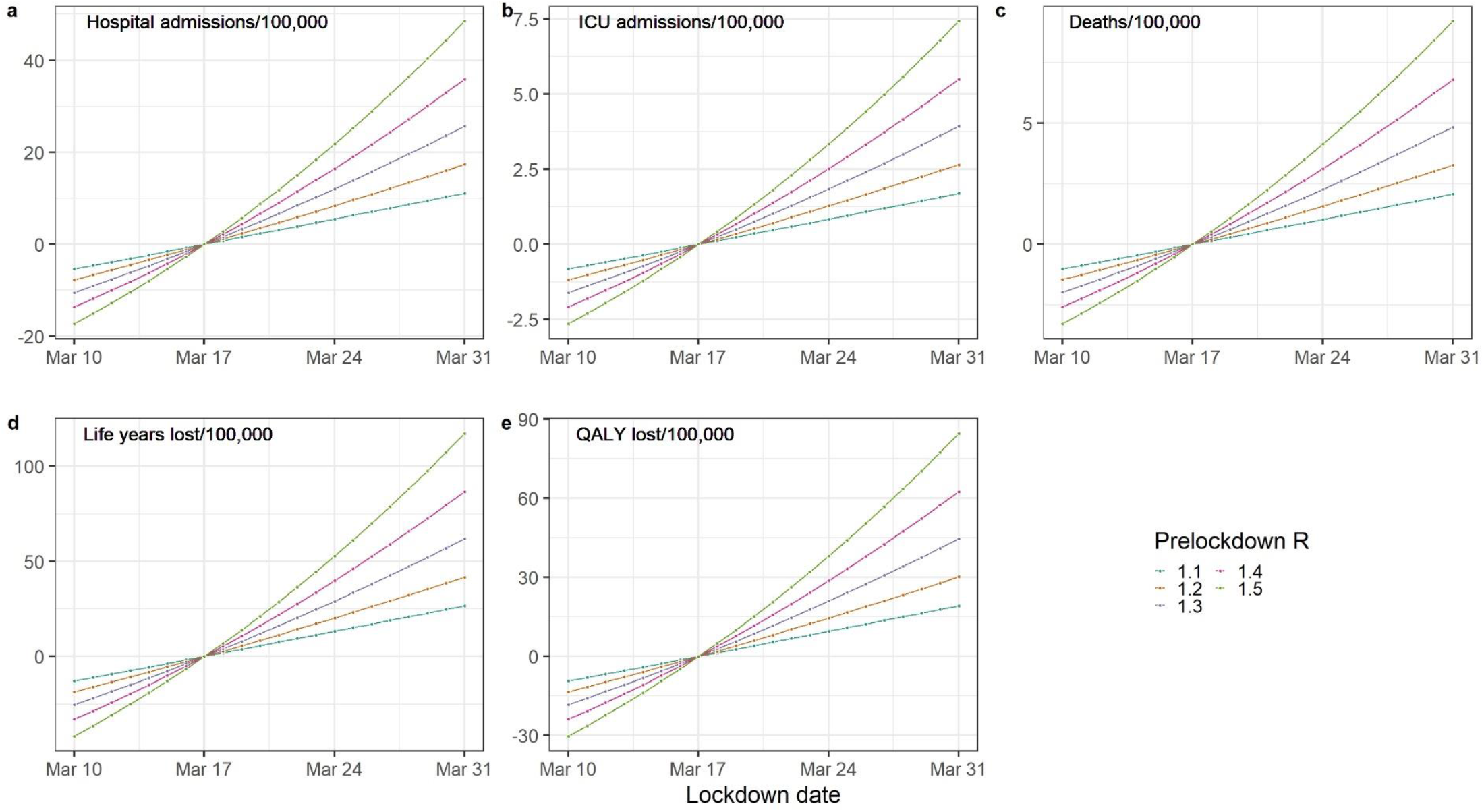
Impact of the change of the lockdown date on the national number of (a) hospital and (b) ICU admissions, (c) deaths, (d) life years and (e) quality-adjusted life years lost per 100,000 inhabitants compared to the observed lockdown date according to the pre-lockdown value of the reproduction number R_prelockdown_ ranging from 1.1 to 1.5. ICU: Intensive care unit. QALY: Quality-adjusted life years.

**Supplementary Figure S11.**
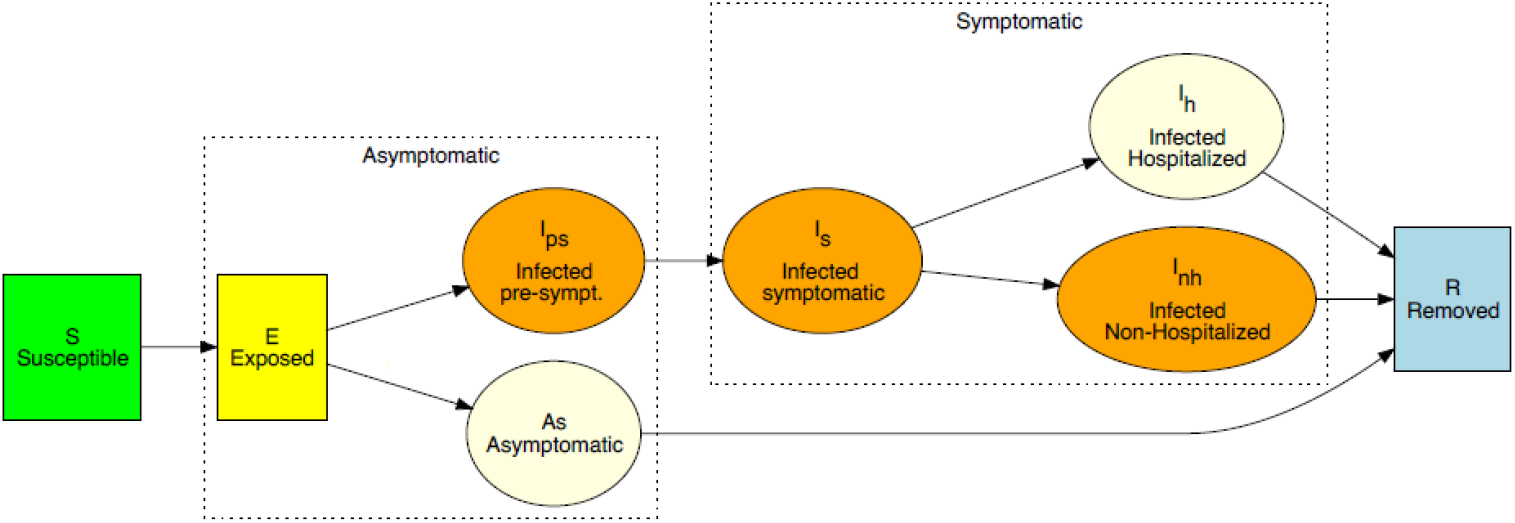
Diagram of the SARS-CoV epidemiological model

**Supplementary Figure S12.**
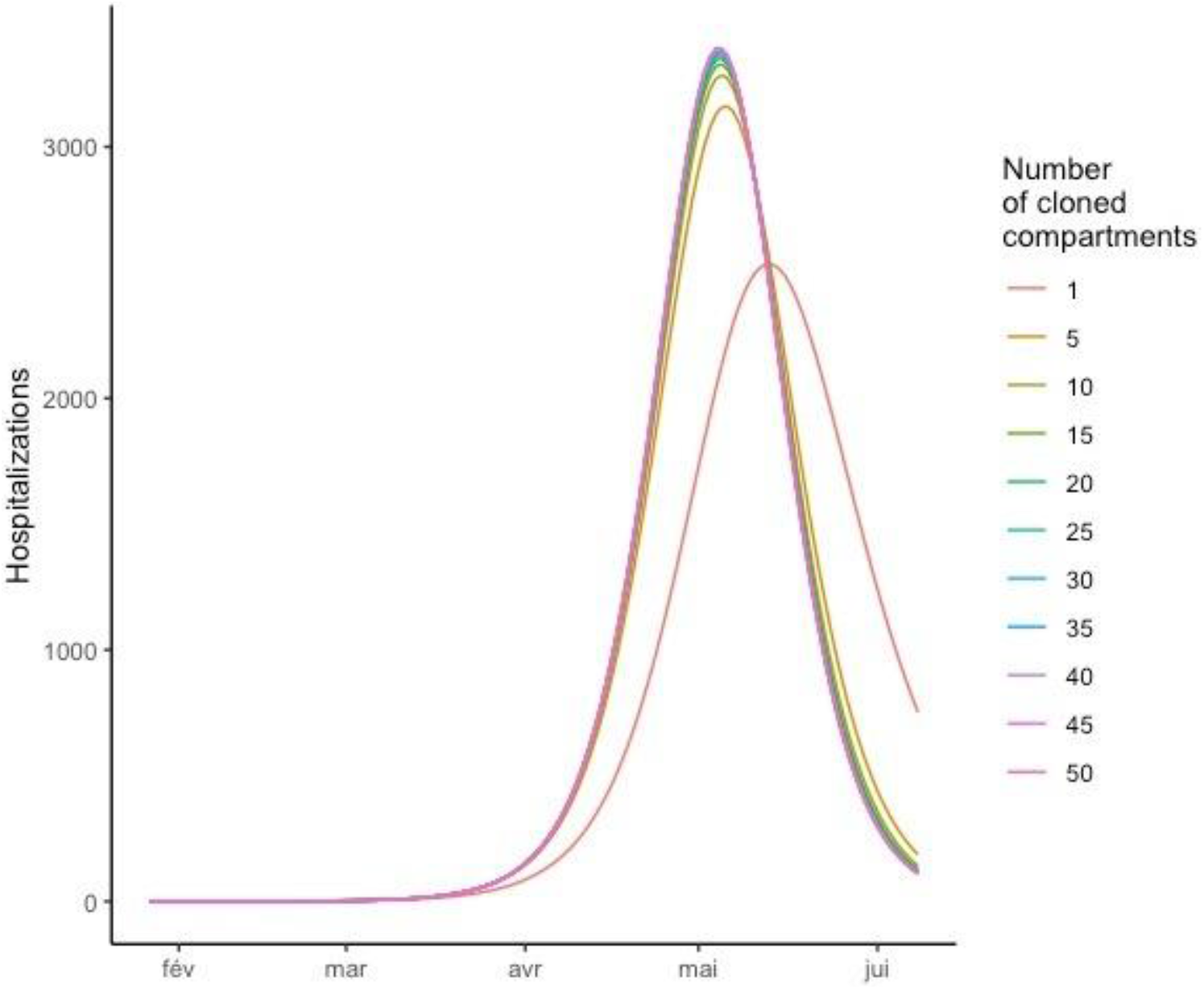
Sensitivity analysis on the number of sub-compartments in each compartment of the transmission model

**Supplementary Figure S13.**
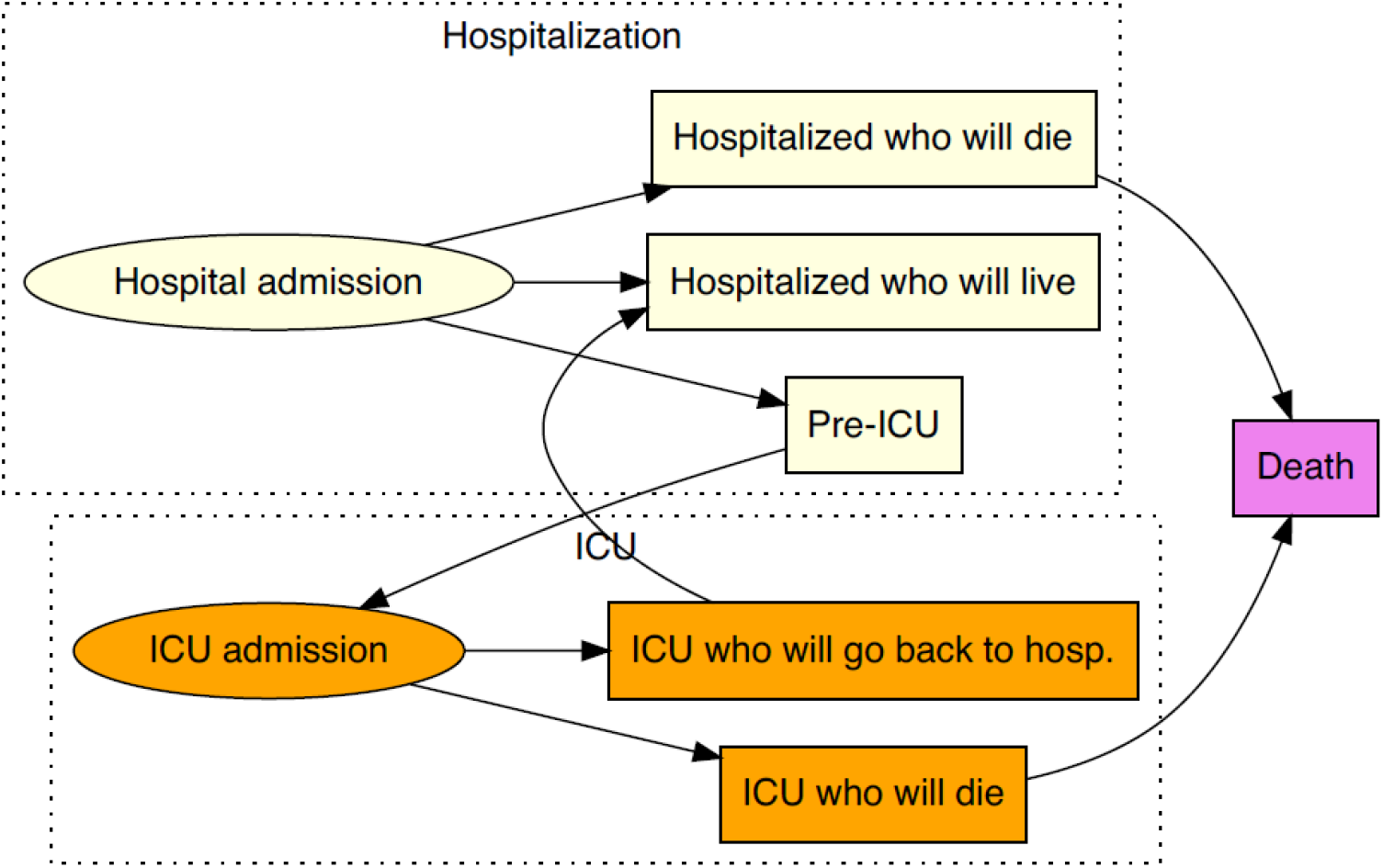
Diagram of the care pathway in hospital settings of a hospitalized COVID-19 infected individual ICU: Intensive care unit.

